# Auditing Large Language Model–Generated Digital Standardized Patients for Demographic Bias: A Simulation Study with HIV Pre-Exposure Prophylaxis Screening as a Tracer Condition

**DOI:** 10.64898/2026.09.01.26361928

**Authors:** Benjamin Daniels

**Affiliations:** McCourt School of Public Policy, Georgetown University, Washington, DC; Department of Global Health and Population, Harvard T.H. Chan School of Public Health, Boston, Massachusetts

## Abstract

Large language models (LLMs) are entering clinical training as digital standardized patients (DSPs), simulated patient encounters the model scripts and portrays. Demographic associations learned from corpus co-occurrences can enter at two points: the written case scripts and the live portrayals improvised in role-plays. We audited both pathways using an HIV pre-exposure prophylaxis (PrEP) screening scenario across six demographic factors (age, gender, marital status, sexual orientation, education, and race or ethnicity) in a 216-cell factorial design. With a generated arm and a template-substituted control arm, we simulated 4,320 conversations with fixed audit questions, measuring three channels: the case scripts, the composite role-play trainees receive, and role-play under identical control cases. Differences concentrated where corpus associations were strongest and reinforced predictable stereotypes. Anal sex appeared in 100% of gay men’s cases, 83% of bisexual men’s and 3% of heterosexual men’s; the six factors explained 32% of the variance in composite sexual risk; and education predicted assigned socioeconomic status (*R̄*^2^ = 0.59). Demographics predicted 5 of the 9 improvised probe responses in composite role-play, and 2 under control cases. These two pathways sometimes had opposite signed effects: the model wrote women with higher alcohol use, but role-played them as drinking less, and the two canceled in composite role-play. An audit of either pathway alone would have obscured both effects. These issues are fixable as they are predictable and consistent. LLM-generated DSPs must have their cases and live portrayals audited as separate objects before deployment.

**Author summary:** Medical schools are beginning to use artificial intelligence chatbots as practice patients: the software writes a patient’s story and then plays that patient in a conversation with a trainee. We asked whether these simulated patients portray people differently depending on their demographic profile. We built 216 patient profiles that differed only in age, gender, marital status, sexual orientation, education, and race, asked a widely used language model to write a case for each one in a scenario about HIV prevention, and then ran thousands of practice conversations with the model playing the patient. The written cases assigned riskier behavior to some groups: nearly every case describing a gay man mentioned anal sex, and a profile’s education level predicted the social class the model invented for it. The live conversations showed a second, separate pattern, sometimes in the opposite direction, so the two distortions could hide each other when combined. A program that checks only the written case, or only the conversation, would miss half the picture. We recommend that training programs audit both before students meet these simulated patients.

## Introduction

LLM-powered digital standardized patients (DSPs) are rapidly entering medical education [1–4], extending earlier computerized virtual-patient formats [5] and offering an on-demand alternative to traditional standardized patient programs [6, 7]. In these programs, trainees can now complete clinical encounters independently at any time with a digital standardized patient whose case history or conversational portrayal are produced by a generative model, based on clinical scenarios scripted by experts [8]. The technical appeal is obvious: more practice repetitions, broader case variation, rapid feedback, and lower cost. But as trainees and providers learn by doing, the cases and their enactments themselves may propagate demographic stereotypes that models encoded from their training data.

Prior studies of LLM bias in medicine have examined the LLM as clinician, for example in clinical decision support systems. This study engages a distinct pathway for bias to enter medical education: the LLM as patient. As in LLM-as-provider studies, we are concerned with the fact that LLMs deeply and invisibly encode social associations between demographic identity and clinical content [9–11]. In clinical tasks, for example, they have previously been shown to produce biased recommendations [12], to propagate race-based medicine [13], and to generate demographically stereotyped clinical vignettes [14]. Racial bias has also been reported in psychiatric diagnosis [15].

In a DSP where the LLM is the patient, biased patient portrayals in training can shape clinical intuitions – case-based pattern recognition built through repeated experiential exposures [16, 17] – before trainees have developed the expertise to question these invisible correlations. Those portrayals then become part of the hidden curriculum [18–20], and they are especially challenging for course designers and trainees to review when such cases are portrayed at scale. After all, any *individual* case is plausible, but bias compounds through subtle accumulations of differential patterns.

To investigate how these mechanisms might work in DSPs, we used a scenario of HIV pre-exposure prophylaxis (PrEP) evaluation and prescribing as a lens to investigate potential issues in the generation and portrayal of simulated patient scenarios. We selected this scenario both because it is likely to reveal the types of patterns LLMs may encode and because it is important in its own right. Reproductive health in general and HIV/AIDS care in particular are high-risk clinical areas where stereotypes already inform care disparities. For example, Black and Latino men who have sex with men bear a disproportionate HIV burden but experience lower PrEP uptake, partly because of provider-side barriers and stigma [21–24].

PrEP screening requires discussing sexual behavior, substance use, and relationship structure – exactly the types of difficult topics through which a training environment that encodes differentially biased information about population risks and behaviors can reinforce demographic assumptions that clinicians should be learning to question.

Contemporary medical education, unfortunately, provides little structured training on the health needs of sexual and gender minorities [25].

Such inappropriate demographic differences can enter a DSP at two points: in the case scripts a model writes before an encounter, as well as through the portrayals it improvises during role-play based on differential demographics even when the underlying clinical facts are “identical”. This would occur in a simulation that attempts to hold clinical features constant while observing or encouraging provider practice across a demographically representative simulated population. To test these channels, we ran a simulation experiment with 4,320 simulated clinical conversations spanning a full factorial of six demographic factors. We adapted the logic of audit studies of clinical care [26, 27] to the model itself. Specifically, from a single base case, we had LLMs (1) generate derivative scripts for members of various demographics; (2) enact these scripts; and (3) enact scripts that had been created for those same demographics in a deterministic fashion (that is, by inserting only the demographic identifiers).

Across these three channels, we found that LLM-generated DSPs imported demographic correlations inappropriately in both generative stages, even when asked to hold clinical factors constant across those demographics. For example, in the generated case libraries, every gay patient practiced anal sex, women consistently presented with fewer sexual and behavioral risk factors, and education mapped near-deterministically to socioeconomic status. These are exactly the types of associations that PrEP training should teach clinicians to question, so that care related to one of the clinical factors is not assumed from the demographic presentation.

We also found that these associations could behave in opposite directions in generation and enactment phases, meaning that a final screen only could miss these flaws. However – reassuringly – we also found that these differential presentations were moderate, concentrated, and largely reproducible across repeated conversations, even in general-purpose models not fine-tuned for clinical simulation. Because they are foreseeable, addressing them may not require expensive technical mitigation like model fine-tuning. The core challenge is then human and social – deciding which of the demographic associations imported by the model are clinically acceptable and which are not.

We therefore ask: when an LLM produces a digital standardized patient, does it introduce demographic differences through the case scripts it writes, through the portrayals it improvises during role-play, or through both? And can educators detect each pathway by reviewing the scripts alone? We proceed in three steps. First, we compare the 216 generated case scripts to measure differences in what the LLM wrote. Second, we examine improvised responses in conversations based on those generated scripts, the composite exposure a trainee would actually receive. Third, we isolate enactment differences by holding case content identical across demographic labels and observing how the portrayal alone changes.

## Conceptual framework

Clinical reasoning develops through case-based pattern recognition. Trainees exposed to repeated exemplars of a presentation build mental models of “illness scripts” linked to patient presentations, knowledge structures linking specific diagnoses to the contexts and patients in which it appears, including the demographic profile of who presents with what [28, 29]. Novices rely on this knowledge base most heavily before obtaining the analytic knowledge needed to override it [16], and experiential learning theory implies that whatever regularities the practice environment contains will be absorbed as priors [17]. The case library a trainee uses for practice therefore informs the statistical distribution from which they infer demographic base rates, and are formed at the stage when the trainee cannot yet distinguish epidemiology from stereotype. Demographic correlations that enter this library unreviewed form the “hidden curriculum”, or clinically relevant content implicit in the structure of the training environment [18, 19].

Language models acquire their associations much the same way. An LLM is trained to reproduce the co-occurrence statistics of its corpus, and demographic terms co-occur with clinical and behavioral content in systematic ways, so the trained model encodes those associations [9–11]. When prompted to script or role-play a persona, the model imagines or enacts a character conditioned on those demographic cues, using its learned associations to fill in whatever the prompt leaves unspecified [30]. That filling-in is what makes generative case writing cheap: a single base scenario can become hundreds of plausible, fully detailed patients quickly, for example with the goal of filling out a population-representative training pool. But such models import stereotyped content by the same process. At *case generation*, the imported associations shape the written script.

Those differences are fixed before any encounter occurs and are in principle reviewable. At *enactment*, they shape the portrayal at run-time, in answers that exist only once a trainee has asked the question, where no advance review can inspect them.

## Methods

In this simulation study, we developed an underlying clinical scenario of PrEP screening and, based on demographic profiles, had an LLM either generate a new script, or we deterministically inserted those identifiers into the base script. Then, the LLM role-played the scripts responding to a set of pre-determined screening questions, the answers to which formed our analysis corpus. The clinical facts of the case were set in advance, and the case involved no diagnostic decisions, so the design estimated whether case content and portrayal varied with demographic assignment while the scenario intended to hold those facts constant [4].

We conducted a full factorial simulation experiment crossing six demographic factors (age, gender, marital status, sexual orientation, education, race/ethnicity) to produce 216 unique patient profiles for the PrEP screening scenario (Figure 1; Table 1). The factorial design assigned the demographic labels, and no label was randomly assigned. Each profile was one cell of the full crossing, so every combination of labels appears exactly once and the six factors were orthogonal by construction. In the *generated arm*, Claude Sonnet 4 (Anthropic) rewrote a base case script for each profile, filling in unspecified clinical details under an explicit instruction not to rely on demographic stereotypes. In the *control arm*, cases were adapted by template substitution with no LLM involvement.

**Figure 1:**
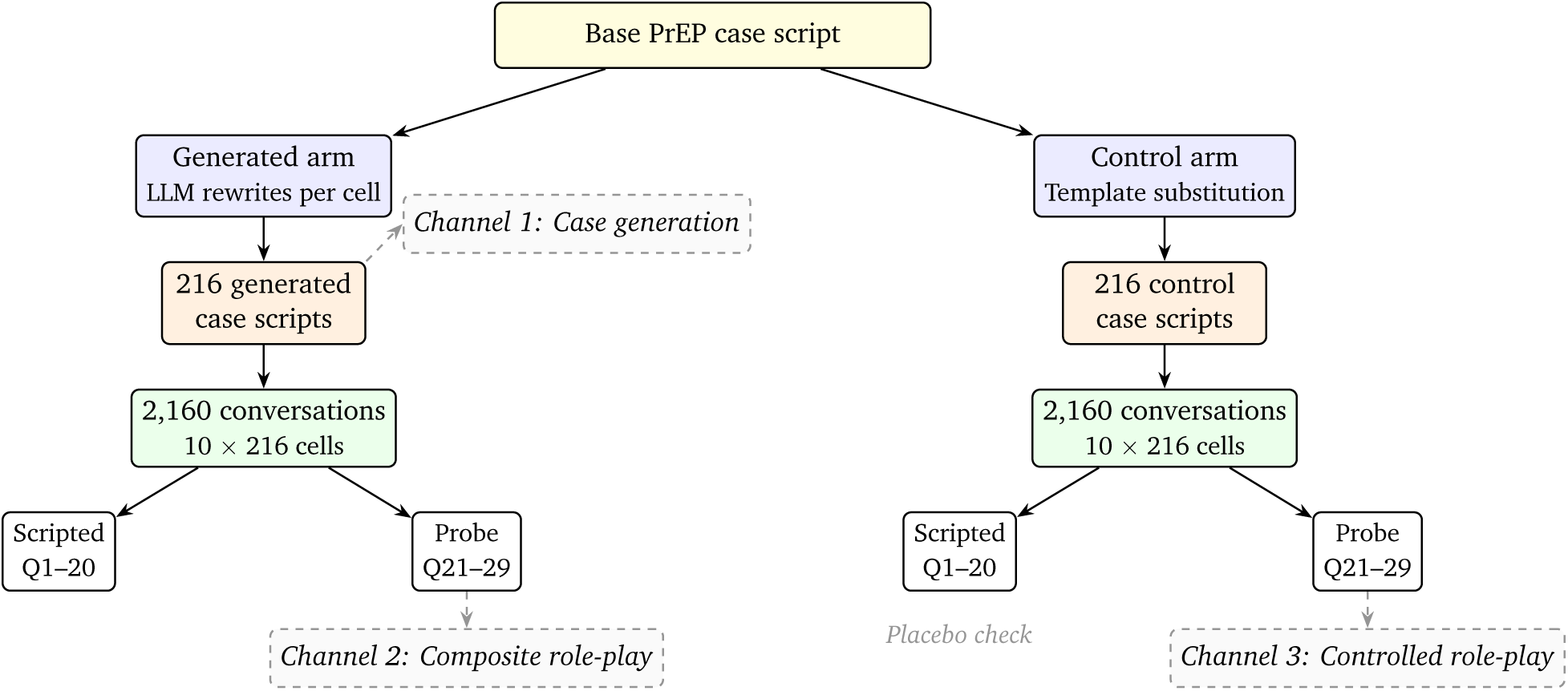
Study design and three-channel decomposition framework. The base case script is adapted via two arms: an LLM-generated arm (left) and a template-substituted control arm (right). Each arm yields 216 case scripts and 2,160 simulated conversations. Questions 1–20 have scripted answers; questions 21–29 (probe questions) are improvised. Channel 1 measures case-generation differences by comparing the 216 generated case scripts. Channel 2 measures composite role-play differences from generated-arm probe responses. Channel 3 isolates role-play differences from control-arm probe responses, where the case content is held identical across demographic cells. Alt text: Flow diagram. A base case script splits into a generated arm and a template-substituted control arm, each producing 216 case scripts and 2,160 simulated conversations with scripted and probe questions, feeding three measurement channels: generated case scripts (channel 1), generated-arm probe responses (channel 2), and control-arm probe responses (channel 3).

**Table 1:** Study design summary.

| Component | Specification | N |
| --- | --- | --- |
| *Factorial design (6 factors, full crossing)* |  |  |
| Age | 25, 35, 45 years | 3 |
| Gender | Man, Woman | 2 |
| Marital status | Single, Married | 2 |
| Sexual orientation | Heterosexual, Homosexual, Bisexual | 3 |
| Education | College, High school | 2 |
| Race/ethnicity | White, African American, Hispanic | 3 |
| **Total cells** |  | **216** |
| *Study arms* |  |  |
| Generated | LLM rewrites base case per cell | 216 cases |
| Control | Template substitution, no LLM | 216 cases |
| *Simulated conversations* |  |  |
| Runs per cell per arm | 10 independent runs | – |
| Scripted questions | Fixed responses in case script (Q1–Q20) | 20 |
| Probe questions | Improvised by patient LLM (Q21–Q29) | 9 |
| **Total conversations** |  | **4,320** |
| *Measurement channels* |  |  |
| Channel 1 | Case generation: generated case content | 216 |
| Channel 2 | Composite role-play: generated-arm probe responses | 2,160 |
| Channel 3 | Controlled role-play: control-arm probe responses | 2,160 |

Variation in subsequent role-play therefore depended on the demographic labels alone, and not on the associated characteristics a generative model could import alongside them [31].

For each cell, a second LLM (GPT-4o-mini, OpenAI) role-played the patient in 10 independent conversations (*N* = 4,320 total). We accessed both models through their standard public-facing commercial APIs, with no enterprise, HIPAA-eligible, or zero-retention configuration, because the instrument handled no real patient data. Both are previous-generation models; but their low cost and wide use mean they remain deployed broadly across production applications, and we intend the interpretations here to indicate the types of biases such models *can* have, not what any model set *will* have. In each conversation the patient LLM answered 20 “scripted” questions with pre-written responses and 9 “probe” questions the script did not cover – the kind of unprompted question that often arises in real encounters.

This yielded three measurement channels: *case generation* (channel 1) compared the 216 case scripts directly; *composite role-play* (channel 2) examined probe responses in the generated arm; *controlled role-play* (channel 3) examined probe responses in the control arm, where identical (non-generated) case scripts built by deterministic text substitution isolated the LLM’s own demographic associations. The three channels separated two questions: “would screening the script catch the difference?” (channel 1) and “does the model improvise differently for different demographics, regardless of the script?” (channel 3).

We extracted clinical characteristics and conversation responses using deterministic keyword matching – no LLM was involved in measurement – and regressed them on all six demographic factors. We reported joint *F* statistics rather than directional coefficients as the primary test. Several factors had three levels, so no single signed coefficient summarized them. The audit question was whether demographics predicted clinical content. Every scenario in the design was meant to be clinically identical, so consistent patterning in any direction was a difference of interest that was imported by the LLM either in the case generation or enactment stage. We also reported Benjamini–Hochberg *q* values, because the design tested many outcome-by-factor combinations, and signed coefficients for the individual comparisons where direction was meaningful.

These 10 conversations in a cell shared a single case script, so the nominal 2,160 conversations per arm overstated the information available. In both panels of Table 2 we reported an effective sample size: *N*_eff_ = *N/*DEFF, where DEFF = 1 + (*n*_cell_ *−* 1) ICC, *n*_cell_ = 10, and ICC was the intraclass correlation across the 216 cells, with negative between-cell variance truncated at zero. All six factors were constant within a cell. For the two role-play channels we therefore took the joint *F* and *q* from the specification fitted to the 216 cell means, and the signed coefficients from the conversation-level fit with standard errors clustered on the cell. We generated one case per cell, so we fitted channel 1 at the case level and reported an adjusted *R*^2^.

**Table 2:**
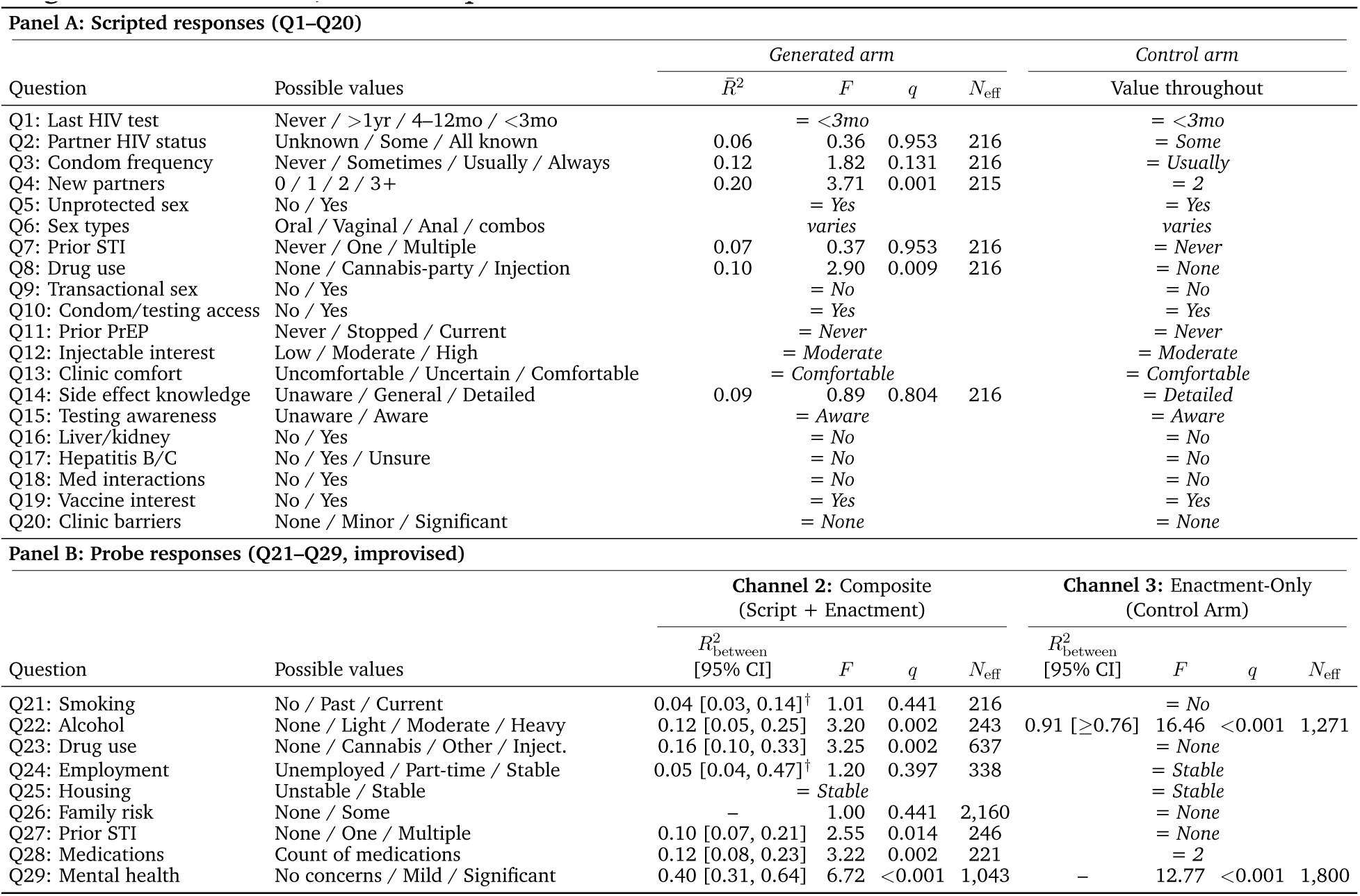
Demographic variation in conversation responses. Unit of observation: the conversation, 2,160 per arm, in 216 factorial cells of 10. Outcomes are regressed on the same six demographic factors as in Table 3 (nine regressor degrees of freedom). Panel A, the responses pre-written into the case script: the adjusted *R̄*^2^, *F* and *q* from one regression, clustered on the 216 cells. Panel B, the probe responses: *R*^2^_between_, the proportional reduction in the between-cell variance component, with a 95% interval from 2,000 cell resamples in brackets; *F* and *q* from a separate fit to the 216 cell means. *q* is Benjamini-Hochberg corrected across outcomes within panel and channel; *N*_eff_ discounts *N* for clustering within a cell. Encodings: *†*, the interval covers the value nine regressors produce by chance; “*≥*”, the upper end is the parameter-space boundary, not a quantile; “–”, *R*^2^_between_ not estimable; “= value”, constant throughout that arm and not fitted; “varies”, no numeric coding. Conversation-level *R*^2^, *F* and *q* are in Table S19; outcomes are on their original coded scales, rows in question order.

The scripted responses in the generated arm did not vary across the 10 runs of a cell. We fitted those at the conversation level with standard errors clustered on the cell, and reported an adjusted *R*^2^ for them as well. The share of variance we reported for the role-play channels was *R*^2^_between_, the proportional reduction in the between-cell variance component of a one-way decomposition of the conversations over the cells, with a 95% interval from a cluster bootstrap over the cells. We reported it in place of the cell-mean *R*^2^ because the cell-mean *R*^2^ depends on the number of runs per cell and the between-cell component does not. Conversation-level *R*^2^, *F* and *q* are in Table S19. Full methods, variable definitions, and prompts are in the online supplementary material.

### Ethics

This study used only large language model–simulated patients and did not involve any human subjects; institutional review board review was therefore not required.

## Results

### Differences in generated case scripts (channel 1)

We extracted 16 clinical characteristics from the 216 generated case scripts (see Method). The model held 4 of them constant regardless of demographics: housing stability, family risk factors, prior STIs, and prior PrEP use. Of the remaining 12, 7 varied with the demographic factors after FDR correction (Table 3). Where the model did not associate a characteristic with particular demographic groups it wrote from a default clinical template, so the differences concentrated on sexual behavior and socioeconomic markers.

**Table 3:** Case generation: demographic variation in all 16 extracted case characteristics (Channel 1). Unit of observation: the generated case, one per factorial cell, 216 in all. *N* falls below 216 where the characteristic could not be extracted from every case document. All three statistics come from one regression of the outcome on the six demographic factors (nine regressor degrees of freedom): the adjusted *R̄*^2^, which can be negative; *F* , their joint test; and *q*, Benjamini-Hochberg corrected within the channel across the fitted outcomes. “Constant” marks a characteristic that took one value and was not fitted; those rows are last, the rest ordered by *R̄*^2^.

| Outcome | Possible values | $N$ | $\bar{R}^2$ | $F$ | $q$ |
| --- | --- | --- | --- | --- | --- |
| SES (implied) | Low / Medium / High | 216 | 0.59 | 35.00 | <0.001 |
| Anal sex mention | No / Yes | 216 | 0.56 | 31.13 | <0.001 |
| Sexual risk (composite) | 0–4 (composite score) | 208 | 0.32 | 11.78 | <0.001 |
| Alcohol use | None / Light / Moderate / Heavy | 216 | 0.28 | 10.52 | <0.001 |
| Sexual partners | Count (1–4) | 208 | 0.17 | 5.76 | <0.001 |
| Condom use | Never / Sometimes / Consistent / Always | 216 | 0.12 | 4.26 | <0.001 |
| Unprotected sex | No / Yes | 216 | 0.08 | 2.96 | 0.004 |
| Drug use | None / Cannabis / Other / Injection | 216 | 0.03 | 1.73 | 0.113 |
| Drug mention | No / Yes | 216 | 0.03 | 1.73 | 0.113 |
| Smoking | No / Past / Current | 216 | 0.00 | 1.01 | 0.481 |
| Employment | Unemployed / Part-time / Stable | 216 | 0.00 | 1.00 | 0.481 |
| Mental health | No concerns / Mild / Significant | 216 | −0.02 | 0.63 | 0.768 |
| Housing stability | Unstable / Stable | 216 |  | *constant* |  |
| Family risk factors | None / Some | 216 |  | *constant* |  |
| Prior STIs | None / One / Multiple | 216 |  | *constant* |  |
| Prior PrEP use | Never / Stopped / Current | 216 |  | *constant* |  |

#### Cultural stereotyping

The model assigned names by race. All 36 African American men were named “Marcus,” Hispanic men received one of two names, and 69% of White men received “Michael” (Table S15). Race or ethnicity predicted 2 of the quantitative outcomes after correction, alcohol use and sexual partners. The three race categories differed on the ordinal alcohol scale (0 = none to 3 = heavy; *F* = 5.22, *q* = 0.026 on the joint test for race), and White patients were assigned higher values than African American patients, the reference level (*β* = +0.15, *q* = 0.129; 95% confidence interval [+0.014, +0.292]). White patients were also assigned more sexual partners (*β* = +0.15, *q* = 0.038).

#### Sexual risk

Sexual risk content varied with the assigned demographics more than any characteristic except socioeconomic status (Table 3). The six demographic factors jointly explained 56% of the variance in whether the case mentioned anal sex (*F* = 31.13, *q <* 0.001) and 32% of the variance in a composite sexual risk score (*F* = 11.78, *q <* 0.001), with orientation and gender the dominant contributors in both. Anal sex content varied with orientation and gender. It appeared in 36 of 36 gay men’s cases, 83% of bisexual men’s, 25% of bisexual women’s, and 3% of heterosexual men’s (Table S12).

Cases generated for women received lower risk scores than cases generated for men (*β* = *−*0.72, *q <* 0.001), with fewer mentions of unprotected sex (*β* = *−*0.09, *q* = 0.007; Table S3). Partner counts did not differ by gender (*β* = *−*0.11, *q* = 0.072). We would have detected a difference of 0.129 partners, or 58% of the composite gender effect. We instructed the model not to rely on demographic stereotypes, and these patterns persisted.

#### Socioeconomic status

Education mapped to occupation and income very strongly (*R̄*^2^ = 0.59; *F* = 35.00, *q <* 0.001; Table S13). College-educated patients were assigned high socioeconomic status in 97% of cases for White patients, 97% for African American patients, and 94% for Hispanic patients, against 17–36% of high-school-educated patients (Table S13). Race differences were smaller and in the same direction: among high-school-educated patients, 36% of White patients received high socioeconomic status, against 17% of African American and 17% of Hispanic patients. Education predicted assigned socioeconomic status more strongly than race did.

### Differences in simulated conversations (channels 2 and 3)

We next asked whether the model also portrayed patients differently once the encounter started. As a manipulation check, we compared the responses the patient LLM gave in the control arm, where we adapted cases by template substitution and not by LLM rewriting, to the answers the base case had scripted (Q1–Q20). Across all 2,160 conversations in that arm, 19 of the 19 numerically coded scripted answers took a single value. The patient LLM reproduced the scripted items exactly, regardless of the demographic label. Any demographic variation in probe responses therefore came from the LLM’s improvisation. A trainee asking an unexpected question often produces the most valuable part of a simulated encounter. In the generated-script arm, by contrast, even the pre-scripted responses showed demographically correlated differences after the case script was re-generated. Married patients were scripted with fewer new partners than single patients (*β* = *−*0.26, *q <* 0.001; Table S5). The gender difference in scripted partner counts was a precise zero (*β* = *−*0.17, *q* = 0.052), and we would have detected 0.150 partners, or 58% of the married contrast. Trainees would encounter the marital-status difference even in the structured portion of the consultation.

Demographic differences appeared in the improvised answers of both role-play channels (Table 2, Panel B). In the generated arm the script and the portrayal moved together, so a difference in a probe answer could have come from the case the model wrote or from the way it then played that case. We separated the two in the control arm, where every cell received the same case content and any difference came from the demographic label and the name alone. Only alcohol and mental health varied there. The other 7 probe items took a single value throughout the control arm and were not fitted, so we could not estimate them in that channel. Both of the differences that remained were by gender.

Demographics predicted 5 of the 9 improvised probe responses across the 216 cells (Table 2). Mental health accounted for the most variance of these (*R*^2^ CI 0.31 to 0.64; *F* = 6.72, *q <* 0.001). The other four were drug use (*R*^2^_between_ = 0.40, 95% = 0.16, 95% CI 0.10 to 0.33; *F* = 3.25, *q* = 0.002), alcohol (*R*^2^_between_ = 0.12, 95% CI 0.05 to 0.25; *F* = 3.20, *q* = 0.002), medications (*R*^2^_between_ = 0.12, 95% CI 0.08 to 0.23; *F* = 3.22, *q* = 0.002), and prior STI (*R*^2^_between_ = 0.10, 95% CI 0.07 to 0.21; *F* = 2.55, *q* = 0.014). At the conversation level, 45-year-olds reported better mental health than 25-year-olds (*β* = *−*0.14, *q* = 0.002), and married patients reported less drug use than single patients (*β* = *−*0.13, *q* = 0.020). Drug use did not differ by gender (*β* = *−*0.09, *q* = 0.164). We would have detected 0.106 of a category, which is 75% of the sexual-orientation difference in drug use in the same channel (Table S25).

Cases in the control arm were identical across all demographics, so any difference in how the patient LLM played a case came from the demographic label and name alone.

Demographics still predicted 2 outcomes: alcohol (*R*^2^_between_ = 0.91, bounded below by 0.76 at 95%; *F* = 16.46, *q <* 0.001) and mental health (*F* = 12.77, *q <* 0.001), whose between-cell component is too small to estimate a share against. Both were gender effects. Women were portrayed as drinking less (*β* = *−*0.12, *q <* 0.001) and reporting worse mental health (*β* = +0.15, *q <* 0.001). White patients were portrayed as drinking more than African American patients (*β* = +0.12, *q <* 0.001; Table S17). Figure 2 plots cell means for six outcomes, two drawn from each channel. Figure 3 reports cell means for all nine probe items in both role-play channels for men and women as a demonstrative example.

**Figure 2:**
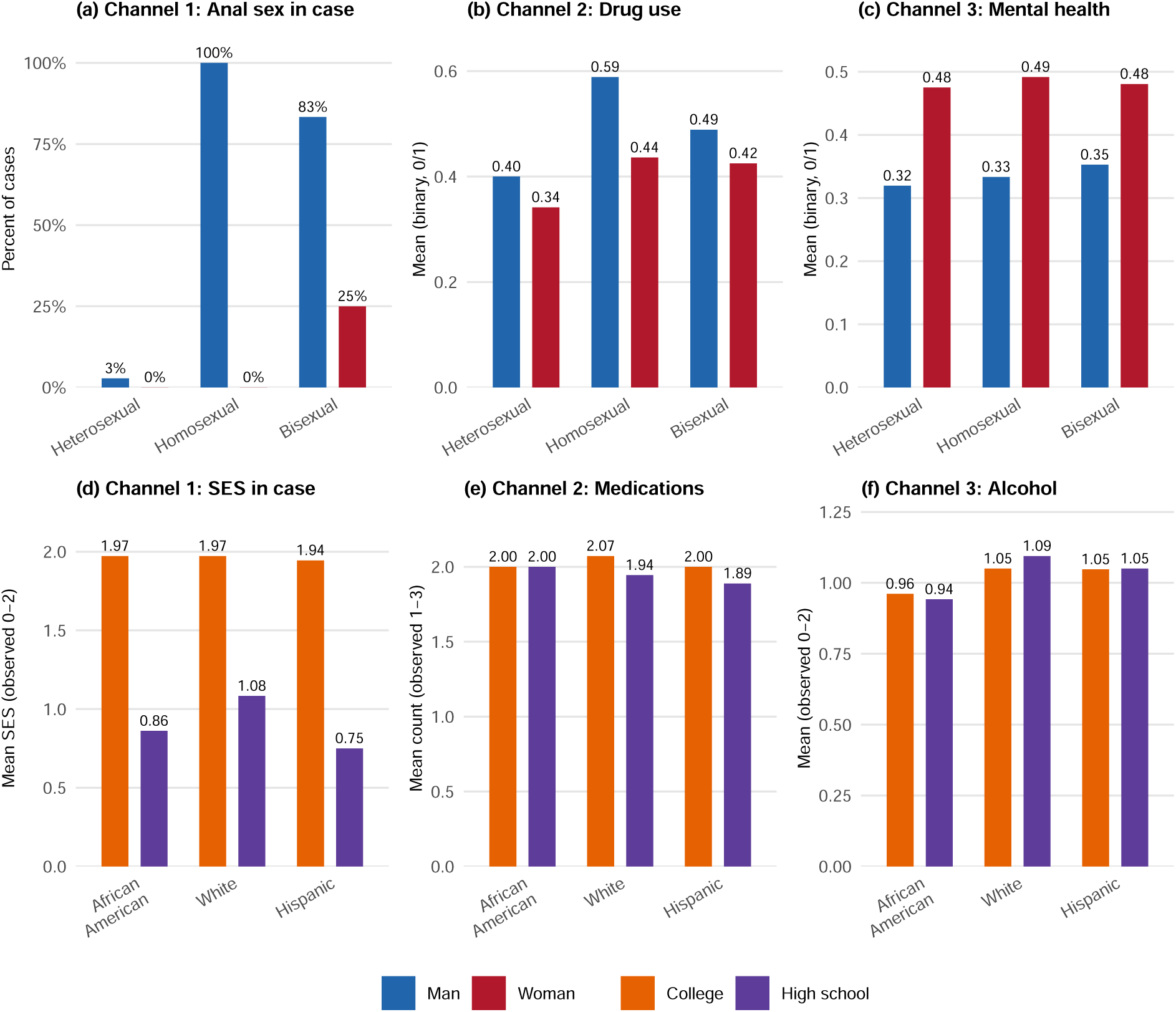
Six outcomes across the three measurement channels. Each bar is a cell mean, printed above the bar. Panels (a) to (c) group cells by sexual orientation and gender; panels (d) to (f) group them by race/ethnicity and education. Panels (a) and (d) are read from the generated case scripts (channel 1), at 36 cases per bar. Panels (b) and (e) are improvised probe responses in the generated arm (channel 2), and panels (c) and (f) are improvised probe responses in the control arm (channel 3), at 360 conversations per bar. Panel (a) plots the percentage of cases that mention anal sex. The other five plot means of coded items: drug use as none, cannabis, other, injection; mental health as no concerns, mild, significant; implied SES as low, medium, high; alcohol as none, light, moderate, heavy; and medications as a count. Alt text: Six panels of grouped bar charts showing cell means for anal sex mention, drug use, mental health, implied socioeconomic status, alcohol use, and medication count, with cells grouped by sexual orientation and gender in the top row and by race/ethnicity and education in the bottom row, compared across the three measurement channels.

**Figure 3:**
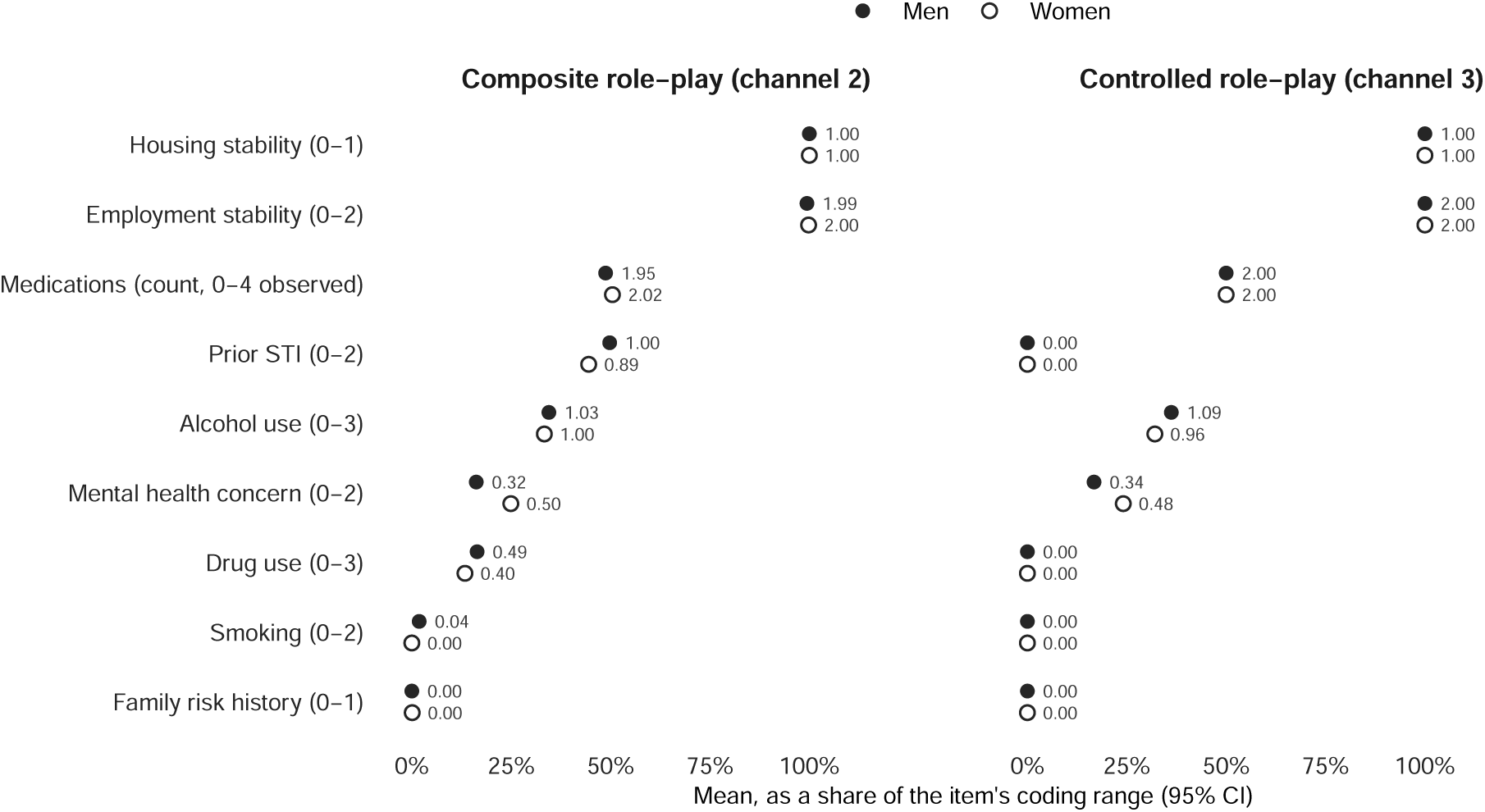
Nine risk-factor items, men against women, in both role-play channels. The probe instrument asks the role-played patient about nine risk factors, so these items exist in the two role-play channels and nowhere else: composite role-play, in which the case script is itself model-generated, and controlled role-play, in which one template case is substituted into each demographic cell. Each point is a cell mean over 1,080 conversations. The items run on scales of different length, so a point is placed at the mean as a share of that item’s coding range; the raw mean is printed beside the point and the coding range is named in the item label. Bars are 95% confidence intervals, narrower than the plotting symbol for most items. Items are ordered by their composite role-play mean. The control template fixes drug content, so controlled role-play is a structural zero in all 2,160 control conversations. Alt text: Dot plot of nine risk-factor items in two panels, composite role-play on the left and controlled role-play on the right, with filled circles for men and open circles for women placed at each item’s mean as a share of its coding range, raw means printed beside each point, and 95% confidence interval bars.

### Cross-channel comparison of generation and enactment effects

Differences in case writing and differences in role-play sometimes ran in opposite directions (Figure 4; Table S18), so a program auditing one channel alone would have overstated one difference and missed the other. The two did not simply compound.

**Figure 4:**
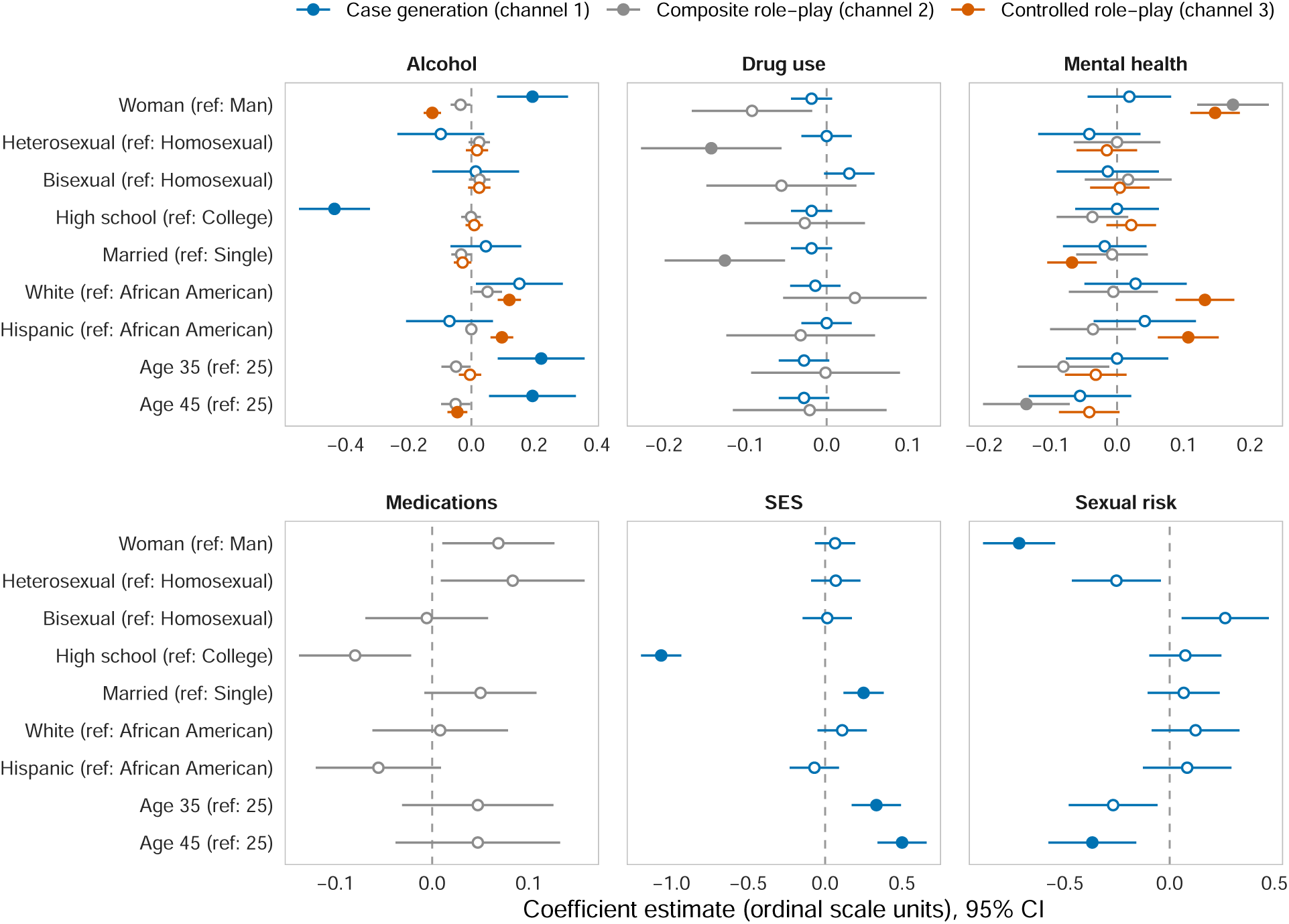
Demographic effects by measurement channel. Points are OLS coefficients with 95% confidence intervals from regressions of each outcome on all six demographic factors; filled points are significant after Benjamini–Hochberg correction (*q <* 0.05), open points are not. Case generation (channel 1, blue) and controlled role-play (channel 3, orange) are the two pathways the framework distinguishes; composite role-play (channel 2, gray) is the mixture a trainee would encounter. SES and sexual risk are case-content outcomes and exist only in channel 1. Alt text: Coefficient plot of demographic effects on eight outcomes, with points for OLS coefficients and horizontal 95% confidence interval bars, colored by measurement channel (case generation in blue, composite role-play in gray, controlled role-play in orange), filled where significant after Benjamini–Hochberg correction and open where not.

The two channels enacted opposite signed effects for alcohol use by gender. LLM case generation gave women *higher* alcohol use (*β* = +0.19, *q* = 0.007; channel 1), and controlled role-play portrayed them as drinking *less* (*β* = *−*0.12, *q <* 0.001; channel 3; Table S18). We estimated the difference between the two channels directly in a stacked regression (interaction *β* = *−*0.32, *p <* 0.001; Table S24). The two effects canceled in channel 2, where the gender effect on alcohol was a precise zero (*β* = *−*0.03, *q* = 0.196; Table S18). We would have detected 0.045 of a category there, which is 23% of the generation effect being canceled (Table S25), so a difference the size of either component would have appeared in channel 2. An analysis sampling only generated-arm encounters would conclude that the model produced no alcohol-related gender difference, when two opposing differences were both present.

Drug use varied with demographics in composite role-play (*R*^2^_between_ = 0.16, 95% CI 0.10 to 0.33; *F* = 3.25, *q* = 0.002). Married patients reported less use than single patients (*β* = *−*0.13, *q* = 0.020), while the gender difference was a precise zero (*β* = *−*0.09, *q* = 0.164). We could not decompose this outcome across channels. The control template fixed drug content, so drug use did not vary in controlled role-play, and in case generation the joint test was *F* = 1.73, *q* = 0.113 (Table S1). The composite difference therefore did not come from explicit drug content in the script. Occupation and lifestyle framing, neither of them clinically relevant, varied across generated cases.

Mental health behaved the same way in both role-play channels. Women were portrayed with worse mental health in composite and in controlled role-play alike (*β* = +0.15, *q <* 0.001 in channel 3), regardless of case content. In the case scripts we would have detected an effect of 0.090, or 52% of the enactment effect (Table S25).

## Discussion

Consider a medical school using LLM-generated DSPs for PrEP counseling training. Its trainees would encounter a case library in which every gay man practices anal sex, education determines socioeconomic status, and women are written with less sexual risk than men. They would then meet a patient whom the model played as drinking less and reporting worse mental health when she was a woman, whatever the script said. These patterns were consistent across encounters. Provider biases contribute to PrEP access disparities [21, 22], and a case library like this one could reinforce them. The differences were strongest along sexual orientation, gender and education, the dimensions most consequential for PrEP training and HIV prevention equity, and they persisted even when the prompt explicitly instructed the LLM not to rely on demographic stereotypes.

The existing literature on LLM bias in medicine and medical education studies the model as clinician, asking whether it recommends care differently across patient demographics [12–14]. We asked the same question of the model as simulated patient, which is the role LLMs are now being given as programs build training and evaluation simulations for medical learners. In that role the model occupies two positions at once, case author and role-player. Our contribution is to treat those two positions as separate objects of measurement. This complements work on how LLMs adopt and enact assigned personas [30].

Generation and enactment differences partially canceled here, so the aggregate behavior of a deployed system is a poor guide to either component. The written script and the run-time portrayal draw on the model’s associations at different points and under different instructions, so the same association appeared in one channel with the opposite sign in the other. For alcohol, case generation and controlled role-play gave the gender effect opposite signs, and the two canceled in the composite arm (Table S18).

For now, programs deploying LLM-generated DSPs should treat generated cases as drafts requiring human demographic review before classroom use, paying particular attention to clinically relevant details that inform common stereotypes. They should also sample live conversations between trainees and DSPs and audit them for demographic patterning in the LLM’s improvised responses. Only run-time monitoring addresses the second entry point [32]. The differences were moderate in size. The gender effect on alcohol under controlled role-play, *−*0.12 on a 0–3 scale, shifted female patients about one-eighth of a category toward lower reporting. Over hundreds of training encounters, even modest per-encounter shifts would add up to a measurably different distribution of patient presentations by gender or orientation. Specifying every clinical variable explicitly in the prompt would eliminate generation differences by construction, but it would also defeat the purpose of generative case writing.

### Limitations

Our results are specific to one clinical scenario, two LLMs, and one prompt formulation. Other clinical domains may show different patterns of demographic difference. Three further limitations are important for interpreting these estimates. First, we extracted outcomes by deterministic keyword matching, and we did not measure differences expressed through tone or phrasing. Second, the simulated encounters followed a fixed physician question sequence, and we did not measure any real world outcomes. We therefore make no claim about trainee learning from repeated exposure to these associations. Answering that would require a separate study with human learners and downstream outcome measurement. Third, the models we audited were current as of the first quarter of 2026, and both remain available at the prices we paid. We did not repeat the audit on later releases. Our design establishes that this class of model behavior occurs. It does not estimate the rate at which any particular model produces it.

### Conclusions

We audited an LLM-based digital standardized patient pipeline across 4,320 simulated PrEP encounters and found systematic demographic differences in both the cases the model wrote and the patients it portrayed, with the two pathways sometimes offsetting each other. The differences recurred across repeated encounters and were confined to a small number of outcomes. As LLM-powered digital standardized patients scale into routine medical training, programs should audit case writing and dynamic role-play as separate objects.

## Author contributions

Benjamin Daniels: Conceptualization, Methodology, Software, Formal analysis, Investigation, Data curation, Visualization, Writing (original draft), and Writing (review and editing).

## Data availability

All code, prompts, case templates, and simulated conversation logs are openly available on Zenodo at https://doi.org/10.5281/zenodo.21405080.

## Acknowledgments

The author used Claude Code (Anthropic) as a coding assistant during analysis scripting and manuscript preparation. The large language models under study (Claude Sonnet 4 for case generation and GPT-4o-mini for patient enactment) were used as experimental instruments, not as tools for drafting or interpreting results.

